# Evaluation of Radiology Residents’ Reporting Skills Using Large Language Models: An Observational Study

**DOI:** 10.1101/2024.11.06.24316838

**Authors:** Natsuko Atsukawa, Hiroyuki Tatekawa, Tatsushi Oura, Shu Matsushita, Daisuke Horiuchi, Hirotaka Takita, Yasuhito Mitsuyama, Ayako Omori, Taro Shimono, Yukio Miki, Daiju Ueda

## Abstract

**Background:** Large language models (LLMs) have the potential to objectively evaluate radiology resident reports; however, research on their use for feedback in radiology training and assessment of resident skill development remains limited.

**Purpose:** This study aimed to assess the effectiveness of LLMs in revising radiology reports by comparing them with reports verified by board-certified radiologists and to analyze the progression of resident’s reporting skills over time.

**Materials and methods:** To identify the LLM that best aligned with human radiologists, 100 reports were randomly selected from a total of 7376 reports authored by nine first-year radiology residents. The reports were evaluated based on six criteria: (1) Addition of missing positive findings, (2) Deletion of findings, (3) Addition of negative findings, (4) Correction of the expression of findings, (5) Correction of the diagnosis, and (6) Proposal of additional examinations or treatments. Reports were segmented into four time-based terms, and 900 reports (450 CT and 450 MRI) were randomly chosen from the initial and final terms of the residents’ first year. The revised rates for each criterion were compared between the first and last terms using the Wilcoxon Signed-Rank test.

**Results:** Among the LLMs tested, GPT-4o demonstrated the highest level of agreement with board-certified radiologists. Significant improvements were noted in Criteria 1–3 when comparing reports from the first and last terms (all P < 0.023) using GPT-4o. In contrast, no significant changes were observed for Criteria 4–6. Despite this, all criteria except for Criterion 6 showed progressive enhancement over time.

**Conclusion:** LLMs can effectively provide feedback on commonly corrected areas in radiology reports, enabling residents to objectively identify and improve their weaknesses and monitor their progress. Additionally, LLMs may help reduce the workload of radiologists’ mentors.

## Introduction

Large language models (LLMs) are artificial intelligence systems designed to process and understand large amounts of human language [1,2]. LLMs are capable of recognizing highly complex human texts and generating human-like responses. The advent of LLMs has also had an impact on the medical field [3], with the potential to provide various benefits to the field of radiology [4,5]. Several studies have been published in various areas related to radiology, such as improving the efficiency and content of radiology reporting [6,7], summarizing reports for the general public [8], detecting errors in radiology reports [9], diagnosing tumor staging from reports [10], applications for radiology specialist examinations [11–14], verifying the diagnostic capabilities of LLMs [15–19], and education for residents [20,21].

Feedback from radiologist mentors is essential in radiology reporting education for residents. In writing a radiology report, residents must detect normal and abnormal findings; select the findings to include, describe, and interpret accurately; make final and differential diagnoses; and propose further examinations or treatments based on these detected findings [22]. To improve residents’ reporting skills, residents must receive feedback from supervisors on corrections, reflect on this feedback, and understand their work. However, it is not feasible for radiologists to evaluate thousands of reports written annually and objectively assess specific areas needing improvement. While LLMs can provide objective evaluations of residents’ reports, research on using LLMs to provide feedback in radiology report training is still limited [20,21], and no studies have been conducted on providing targeted feedback on specific report elements or on evaluating the growth of residents’ reporting skills over time.

Therefore, the purpose of this study was to evaluate how radiology residents’ reports were revised using LLMs compared to finalized reports confirmed by board-certified radiologists and to investigate the growth of each resident’s radiology reporting skill over time. Exploring the potential of LLMs in optimizing and streamlining resident education could reduce the burden on radiologist mentors. If such an LLM-based evaluation system for resident reporting is implemented in the future, residents would not only receive feedback from radiologists but also identify and improve their weaknesses with the help of LLMs. Additionally, from an objective perspective, residents could compare themselves with their peers and track their growth over time.

## Methods

The study protocol was approved by the Ethics Committee of our institution (IRB:2023-015), and the requirement for informed consent was waived due to its retrospective design. The STROBE Checklist was adhered to in this study [23]. No patient-identifying information was disclosed to the LLMs.

This retrospective study collected consecutive computed tomography (CT) and magnetic resonance imaging (MRI) reports generated by nine first-year radiology residents from 2020 to 2022. All reports included in this study were reviewed by board-certified radiologists. The study design consisted of two phases: LLM selection with prompt tuning and evaluation of residents’ reporting skills. To prevent any data overlap, no reports were duplicated between the two phases. Figure 1 presents the report-selection process.

**Figure 1.**
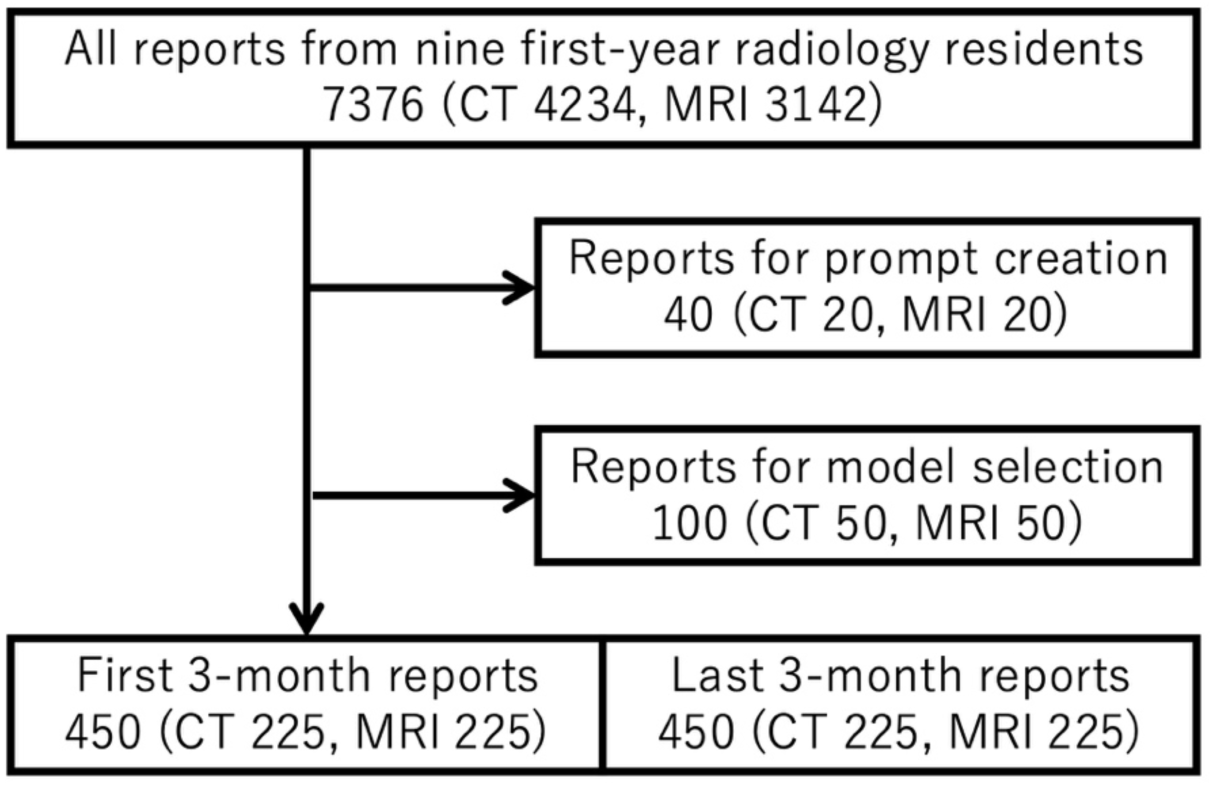
Report selection flow chart illustrating the number of radiology reports included at each stage of the study. The initial pool consisted of 7376 reports (4234 CT and 3142 MRI) from nine first-year radiology residents. Reports were divided into subsets for specific phases: 40 reports (20 CT and 20 MRI) for prompt creation, 100 reports (50 CT and 50 MRI) for model selection, and 450 reports (225 CT and 225 MRI) each for the first and last three-month evaluations of resident reporting skills. This flow chart highlights the distribution and selection process utilized for the retrospective analysis.”

### Evaluation criteria

The following six criteria were assessed by comparing the initial reports written by the residents with the finalized reports confirmed by board-certified radiologists:

***C1. Addition of missing positive findings:*** *Inclusion of findings in the final report that were absent in the initial report, such as newly identified lesions or abnormalities*.

***C2. Deletion of findings:*** *Removal of findings present in the initial report that were deemed normal or non-contributory and therefore excluded in the final report*.

***C3. Addition of negative findings:*** *Explicit mention of the absence of abnormal findings or metastases, especially in response to the primary clinical concern*.

***C4. Correction of the expression of findings:*** *Revisions in the final report addressing inappropriate phrasing, grammatical issues, or inconsistencies present in the initial report*.

*C5. Correction of the interpretation of findings, final diagnosis, or differential diagnosis: Modifications to the interpretation or overall diagnosis in the final report*.

***C6. Proposal of additional tests or treatments:*** *Recommendations for further tests or treatments, such as follow-up MRIs or biopsies, are present in the final report but missing from the initial report*.

Paired initial and finalized reports were input into the LLMs, and the results were evaluated as either positive (1) or negative (0), with a brief rationale provided for each criterion. Figure 2 depicts the prompt used in the LLM evaluation.

**Figure 2:**
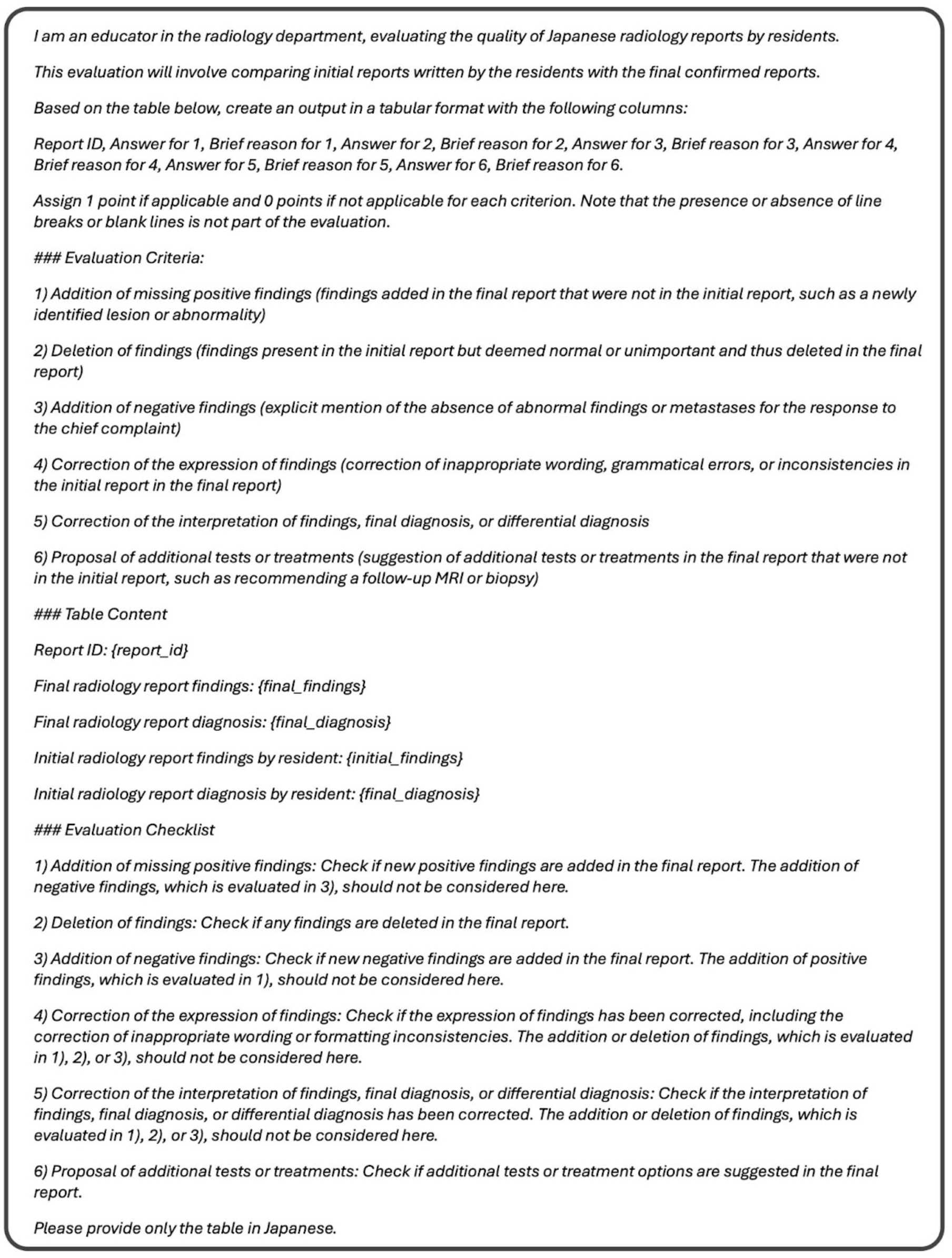
Created prompt for the study used in the evaluation of radiology residents’ reports. The prompt outlines the criteria for assessing the initial radiology reports written by residents against the finalized versions confirmed by board-certified radiologists. It specifies the evaluation process, including six key criteria: (1) addition of missing positive findings, (2) deletion of unimportant findings, (3) addition of negative findings, (4) correction of the expression of findings, (5) correction of the interpretation or diagnosis, and (6) proposal of additional tests or treatments. The prompt format is designed to ensure objective and consistent scoring of report revisions, with 1 point assigned for applicable criteria and 0 for non-applicable items.

### LLM selection with prompt tuning phase

For prompt tuning, 40 reports (20 CT and 20 MRI) were randomly selected to develop the corresponding prompts based on six evaluation criteria. After the prompt-tuning phase, 100 reports (50 CT and 50 MRI) were randomly selected and assessed using the six evaluation criteria by two experienced board-certified radiologists (XX and YY, with 11 and 16 years of experience, respectively) and three LLMs: ChatGPT-4 omni (GPT-4o) [24], Claude-3.5 Sonnet [25], and Claude-3 Opus [25]. During the LLM evaluation, the prompts and CSV files containing the initial and finalized reports were input into the LLMs (GPT-4o API [gpt-4o-2024-08-06], Claude-3.5 Sonnet, and Claude-3 Opus) three times and the most frequently generated answer was determined for each model. Two board-certified radiologists independently reviewed the 100 reports and applied the six evaluation criteria using our facility’s reporting system (SYNAPSE Result Manager; Fujifilm, Tokyo, Japan), where modified lines in the reports are highlighted in different colors for easy visual identification of changes (Supplemental Figure 1). The LLM with the highest agreement rate with the radiologists was subsequently used in the final phase of the study.

### Evaluation of resident’s reporting phase

To evaluate residents’ reporting skills, reports were stratified into four 3-month terms spanning the first academic year: April–June (first term), July–September, October–December, and January–March (last term). Fifty reports (25 CT and 25 MRI) from each of the nine residents were randomly selected from the first and last terms to capture periods that would likely show the most significant improvement in reporting skills for first-year residents. Using the highest-performing LLM, the study analyzed 900 reports with the six evaluation criteria (450 from the first term and 450 from the last term, comprising 225 CT and 225 MRI reports) to assess the extent of revisions made to the reviewed items for each resident.

### Statistical analysis

For the model selection phase, kappa coefficients were calculated to assess the degree of agreement between the radiologists and between either radiologist and the LLMs (GPT-4o, Claude-3.5 Sonnet, and Claude-3 Opus) across the six criteria. The LLM with the highest kappa coefficients, evaluated using the Mann-Whitney U test, was selected to assess the degree of improvement in residents’ reporting skills over time.

For the residents’ report evaluation phase, modification rates for each criterion were calculated for the reports from the first and last terms, with results presented in radar charts. Differences between the first and last-term reports were analyzed using Wilcoxon Signed-Rank tests.

Statistical significance was defined as P < 0.05. Statistical analyses were conducted using Python version 3.9.6 (Python Software Foundation, Wilmington, DE, USA). Kappa coefficient interpretations were as follows: 0–0.20 (slight agreement), 0.21–0.40 (fair agreement), 0.41–0.60 (moderate agreement), 0.61–0.80 (substantial agreement), and 0.81–1.00 (almost perfect agreement).

## Results

The total number of reports included in the study was 7376 (CT: 4234; MRI: 3142), authored by nine first-year radiology residents and confirmed by 23 board-certified radiologists. Table 1 outlines the distribution of reports used for prompt tuning and creation (40 reports), LLM selection (100 reports), and the evaluation of residents’ reporting skills (900 reports).

**Table 1.** Selected reports’ demographics.

| Table 1 Selected reports' demographics |  |  |  |  |
| --- | --- | --- | --- | --- |
|  |  | Number | Mean age | Sex (female) |
| Prompt tuning/creation | Total | 40 | 61 | 19 |
|  | CT | 20 | 64 | 8 |
|  | MRI | 20 | 59 | 11 |
| Model selection | Total | 100 | 58 | 65 |
|  | CT | 50 | 63 | 28 |
|  | MRI | 50 | 54 | 37 |
| Resident evaluation (first term) | Total | 450 | 62 | 229 |
|  | CT | 225 | 65 | 100 |
|  | MRI | 225 | 59 | 129 |
| Resident evaluation (last term) | Total | 450 | 62 | 247 |
|  | CT | 225 | 64 | 110 |
|  | MRI | 225 | 60 | 137 |

### Model selection phase

Table 2 presents the kappa coefficients and accuracy scores for the evaluation criteria (C1–C6) when comparing the two radiologists and the LLMs (GPT-4o, Claude-3.5 Sonnet, and Claude-3 Opus). High inter-rater agreement was observed between the radiologists for all criteria, with kappa coefficients ranging from 0.84 to 1.00. Although the agreement between GPT-4o and the radiologists was somewhat lower than that between the radiologists themselves, it remained substantial for C1, C2, and C6 (kappa coefficients ranging from 0.67 to 0.74; accuracy scores between 0.85 and 0.98). In contrast, lower agreement was noted for C3, C4, and C5 (kappa coefficients between 0.29 and 0.43; accuracy scores between 0.69 and 0.84). GPT-4o, which showed the highest agreement with both radiologists (kappa coefficients for comparisons with radiologist 1: GPT-4o vs. Claud-3.5 Sonnet, P = 0.026; GPT-4o vs. Claud-3 Opus, P = 0.002; and with radiologist 2: GPT-4o vs. Claud-3.5 Sonnet, P = 0.013; GPT-4o vs. Claud-3 Opus, P = 0.002), was chosen for the subsequent evaluation of subsequent residents’ reporting skills.

**Table 2.** Kappa coefficient and accuracy between radiologists and large language models.

|  |  | Criteria 1 |  | Criteria 2 |  | Criteria 3 |  | Criteria 4 |  | Criteria 5 |  | Criteria 6 |  |
| --- | --- | --- | --- | --- | --- | --- | --- | --- | --- | --- | --- | --- | --- |
| Radiologist 1 | vs. Radiologist 2 | 0.90 |  | 0.93 |  | 0.90 |  | 0.92 |  | 0.84 |  | 1 |  |
|  | vs. GPT-4o | 0.72 | (0.86) | 0.67 | (0.89) | 0.40 | (0.73) | 0.41 | (0.69) | 0.29 | (0.80) | 0.74 | (0.98) |
|  | vs. Claud 3.5 Sonnet | 0.15 | (0.56) | -0.04 | (0.81) | -0.15 | (0.60) | 0.51 | (0.77) | 0.39 | (0.75) | 0.26 | (0.91) |
|  | vs. Claude 3 Opus | -0.23 | (0.39) | 0.07 | (0.58) | -0.16 | (0.43) | 0.11 | (0.50) | 0.06 | (0.73) | -0.02 | (0.94) |
| Radiologist 2 | vs. GPT-4o | 0.70 | (0.85) | 0.67 | (0.89) | 0.43 | (0.75) | 0.42 | (0.69) | 0.43 | (0.84) | 0.74 | (0.98) |
|  | vs. Claud 3.5 Sonnet | 0.14 | (0.55) | -0.04 | (0.81) | -0.13 | (0.64) | 0.50 | (0.77) | 0.39 | (0.75) | 0.26 | (0.91) |
|  | vs. Claude 3 Opus | -0.21 | (0.40) | 0.03 | (0.56) | -0.13 | (0.45) | 0.10 | (0.48) | 0.13 | (0.75) | -0.02 | (0.94) |

Values represent the kappa coefficients, with accuracy in parentheses.

### Residents’ reporting evaluation phase

The comparison of the six criteria (C1–C6) in radiology reports authored by nine first-year residents between the first and last terms revealed significant improvement in C1, C2, and C3, while C4, C5, and C6 did not show significant changes (Table 3). However, C4 and C5 displayed trends of improvement over time. Figure 3 illustrates the changes in individual evaluations through radar charts. Overall, enhancements in residents’ reporting were evident from the first to the last term. Notably, residents 2, 4, 5, and 8 demonstrated substantial improvements, while residents 1, 2, 4, and 9 showed only minimal advancements. Resident 3 exhibited partial improvement. Detailed results for CT-only and MRI-only evaluations are provided in Supplemental Table 1 and Supplemental Figures 1 and 2.

**Figure 3:**
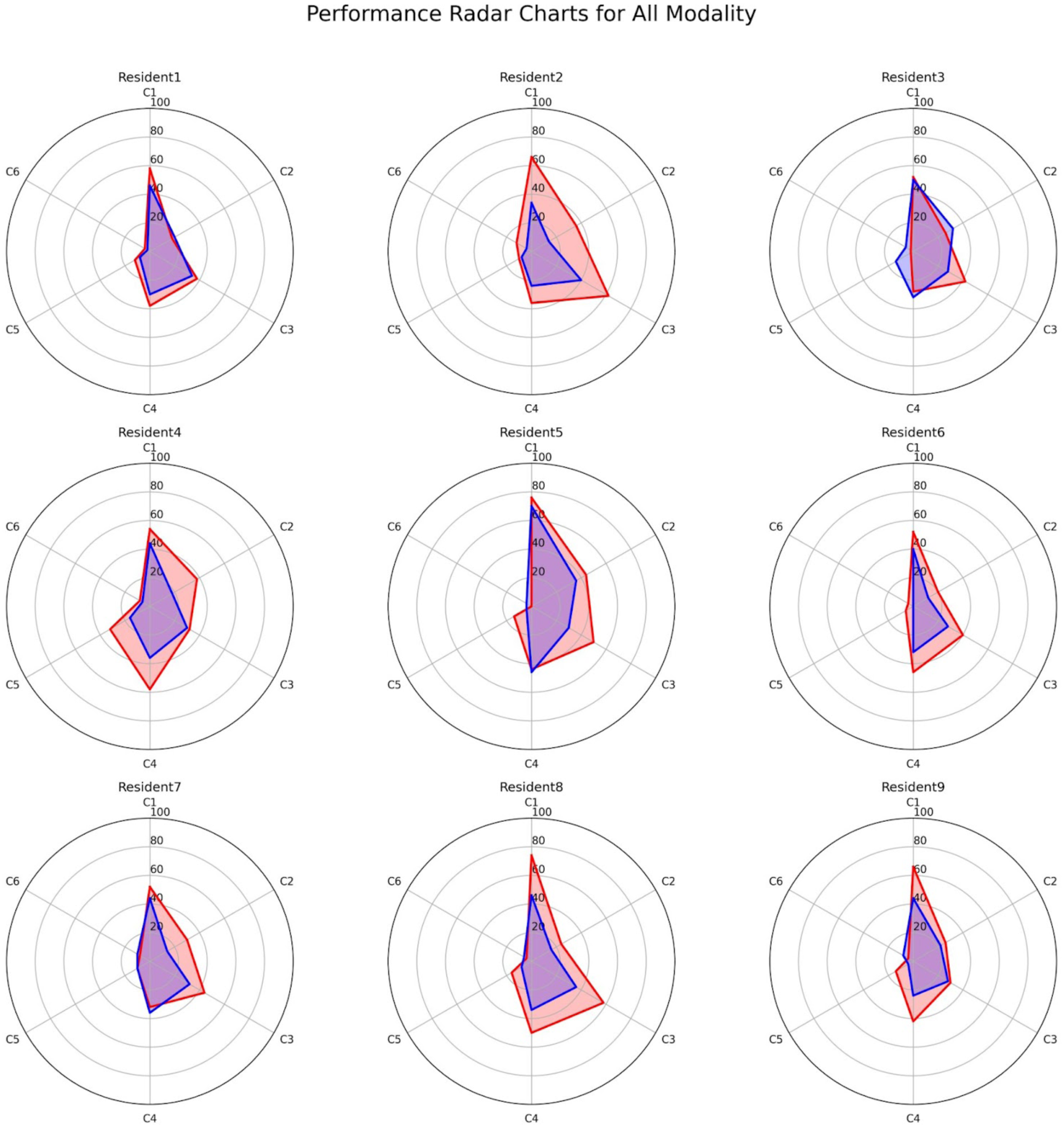
Radar chart showing the modification rates for each criterion (C1–6) among nine first-year residents. Red represents modification rates for the first term, while blue indicates those for the last term. Criteria 1. Addition of missing positive findings; Criteria 2. Deletion of findings; Criteria 3. Addition of negative findings; Criteria 4. Correction of the expression of findings; Criteria 5. Correction of the interpretation of findings; Criteria 6. Proposal of additional tests or treatments

**Table 3.** Revised rate of first-year resident reports.

| Table 3 Revised rate of first-year resident reports |  |  |  |
| --- | --- | --- | --- |
|  | First term | Last term | P value* |
| Criteria 1 | 61% | 46% | < 0.001 |
| Criteria 2 | 29% | 20% | 0.023 |
| Criteria 3 | 44% | 32% | 0.004 |
| Criteria 4 | 42% | 33% | 0.05 |
| Criteria 5 | 13% | 8% | 0.14 |
| Criteria 6 | 5% | 5% | 0.91 |

Data showing mean percentage
\*Wilcoxon Signed-Rank test
Criteria 1. Addition of missing positive findings; Criteria 2. Deletion of findings; Criteria 3. Addition of negative findings; Criteria 4. Correction of the expression of findings; Criteria 5. Correction of the interpretation of findings; Criteria 6. Proposal of additional tests or treatments

## Discussion

This study evaluated the revisions of radiology resident reports by comparing them with reports confirmed by board-certified radiologists using LLMs. GPT-4o exhibited the highest agreement with human radiologists and was selected to assess the progression of residents’ reporting skills. Significant improvements were noted in C1–C3, highlighting the addition of missing positive or negative findings and the deletion of findings between the first and last-term reports.

This study demonstrates that various criteria in radiology reports can be objectively evaluated using LLMs. By employing LLMs such as GPT-4o, residents can receive feedback on revised items, enabling them to objectively identify and improve their weaknesses. A prior study focusing on missed diagnoses in resident reports indicated that LLM-based feedback was cautiously but positively received by residents [20], suggesting that such feedback could enhance their satisfaction with educational processes. Although only six criteria were evaluated in this study, the framework could be adapted by other institutions to incorporate additional criteria for more comprehensive feedback. Furthermore, by facilitating feedback for radiology trainees, LLMs may help alleviate the workload of busy radiologists.

The kappa coefficients between GPT-4o and radiologists were substantial for C1, C2, and C6 (“C1. Addition of missing positive findings,” “C2. Deletion of findings,” and “C6. Proposal for additional examinations or treatments”). This suggests that GPT-4o performed well in identifying the addition or deletion of findings and new recommendations, areas that do not demand complex interpretation. In contrast, the agreement rates for C3, C4, and C5 were fair to moderate. In C4 and C5, which involved revisions of expressions or interpretations of findings and diagnoses, GPT-4o encountered greater challenges in comprehending and accurately evaluating the revised text compared to human evaluators. Previous research has shown that LLMs may overestimate the importance of detected revisions, potentially due to difficulties in understanding the nuanced modifiers and context applied by expert radiologists [21].

Growth rates varied across evaluation items. Significant improvements were noted in C1–C3 (“C1: Addition of missing positive findings,” “C2: Deletion of findings,” and “C3: Addition of negative findings”) between the first and last terms. The improvement in C1 suggests residents may find this criterion more straightforward to enhance over a relatively short period by studying imaging findings and anatomical structures and receiving targeted feedback on commonly missed elements. Similarly, improvements in C2 and C3 were likely driven by a deeper understanding of normal and abnormal images and recognizing the importance of mentioning negative findings to address clinical context, resulting in more accurate reporting. In contrast, while C4 and C5 (“C4: Correction of the expression of findings” and “C5: Correction of the diagnosis”) showed some improvement, changes were not statistically significant. For C4, producing a well-phrased report may be influenced by variable guidelines and subjective interpretations by attending radiologists. Previous studies on resident development have shown that scores for image-based questions tend to improve more rapidly and achieve higher levels than knowledge-based questions [26]. This finding implies that making accurate interpretations and diagnostic conclusions based on imaging may be more challenging for residents than identifying abnormalities, potentially explaining the significant growth in C1–C3 relative to C4 and C5.

Growth rate differences were also noted among the nine residents. Residents 2, 4, 5, and 8 showed marked improvement, with their radar charts from the first term indicating larger areas than others, suggesting a higher tendency for revisions. This pattern implies that greater improvements by the last term might reflect a higher number of initial errors at the start of residency.

This study has several limitations. First, not all residents completed their residency at the same institution, limiting the ability to evaluate consistent improvements in report writing skills. Second, even when the same criterion was revised multiple times within a single report, it was counted as a single revision, potentially causing minor discrepancies between LLM-based evaluations and actual reporting skills. Third, the six evaluation criteria used may not have been sufficient to fully assess resident reports. However, adding or customizing criteria could allow for broader evaluations using these methods. Fourth, this study did not apply fine-tuning to the LLMs. Future advancements in LLMs with appropriate fine-tuning could enable more accurate evaluations.

In conclusion, LLMs can be used to assess revisions in radiology residents’ reports and track their reporting skill development over time. They offer potential feedback on frequently revised items, such as the addition or deletion of findings, helping residents objectively identify and address their weaknesses. LLMs could play a valuable role in resident education by providing feedback and reducing the workload of radiologists.

## Data Availability

Data supporting the findings of this study are available upon request from the corresponding author.

## Supplemental Data

**Supplemental Table 1.** Revised rate of first-year resident reports of CT and MRI.

|  |  | First term | Last term | P value* |
| --- | --- | --- | --- | --- |
| CT | Criteria 1 | 64% | 48% | 0.008 |
|  | Criteria 2 | 31% | 20% | 0.016 |
|  | Criteria 3 | 52% | 39% | 0.008 |
|  | Criteria 4 | 41% | 34% | 0.27 |
|  | Criteria 5 | 13% | 8% | 0.16 |
|  | Criteria 6 | 6% | 7% | 0.7 |
| MRI | Criteria 1 | 59% | 45% | 0.008 |
|  | Criteria 2 | 28% | 21% | 0.22 |
|  | Criteria 3 | 36% | 25% | 0.06 |
|  | Criteria 4 | 41% | 32% | 0.06 |
|  | Criteria 5 | 13% | 8% | 0.19 |
|  | Criteria 6 | 4% | 3% | 0.75 |

Data showing mean percentage
\*Wilcoxon Signed-Rank test
Criteria 1. Addition of missing positive findings; Criteria 2. Deletion of findings; Criteria 3. Addition of negative findings; Criteria 4. Correction of the expression of findings; Criteria 5. Correction of the interpretation of findings; Criteria 6. Proposal of additional tests or treatments

**Supplemental Figure 1.**
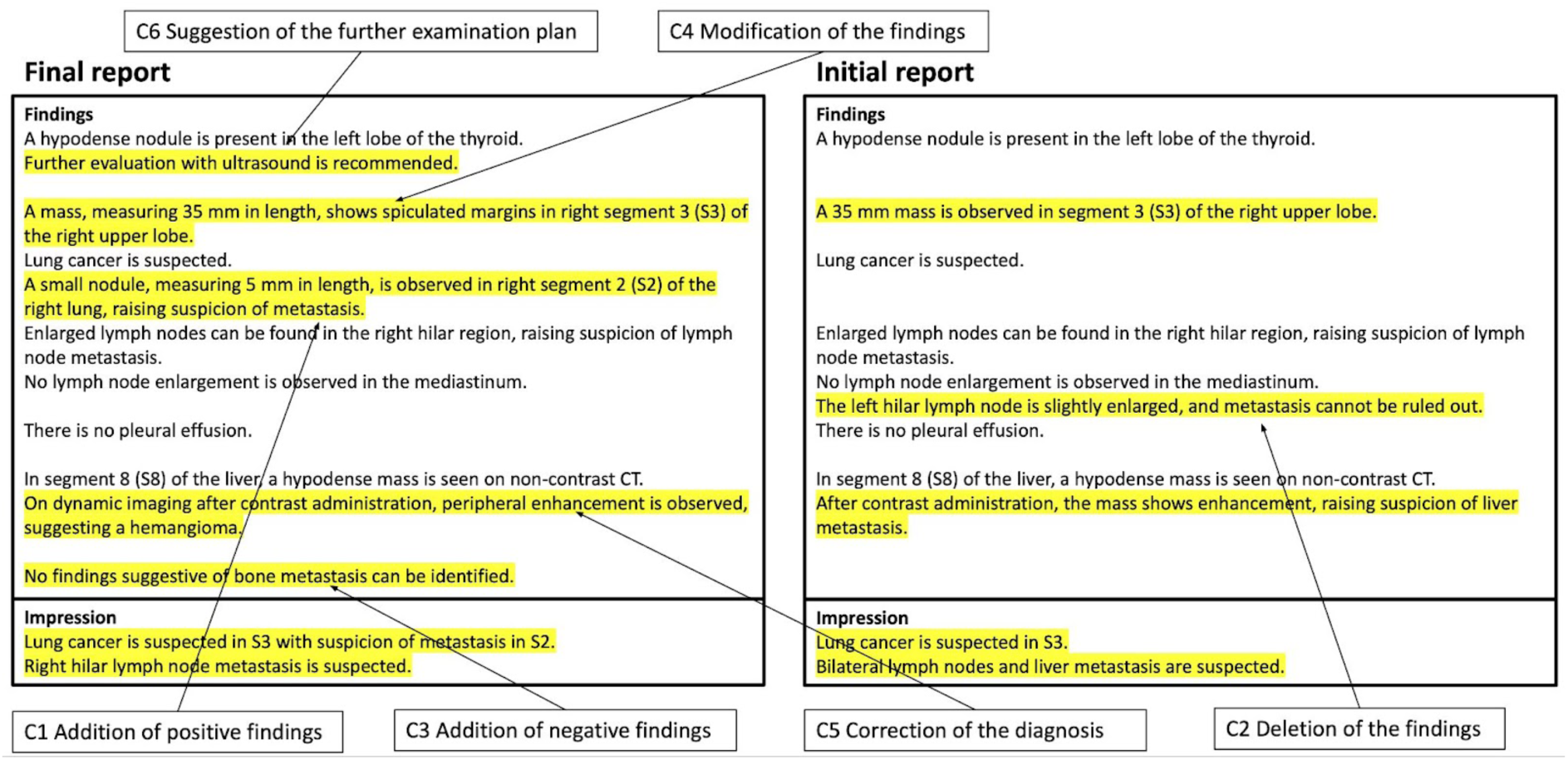
Reporting system used in this study, in which modified lines in the reports are highlighted in different colors, allowing easy visual recognition of changes. An example of criteria 1–6 is shown in C1–6.

**Supplemental Figure 2.**
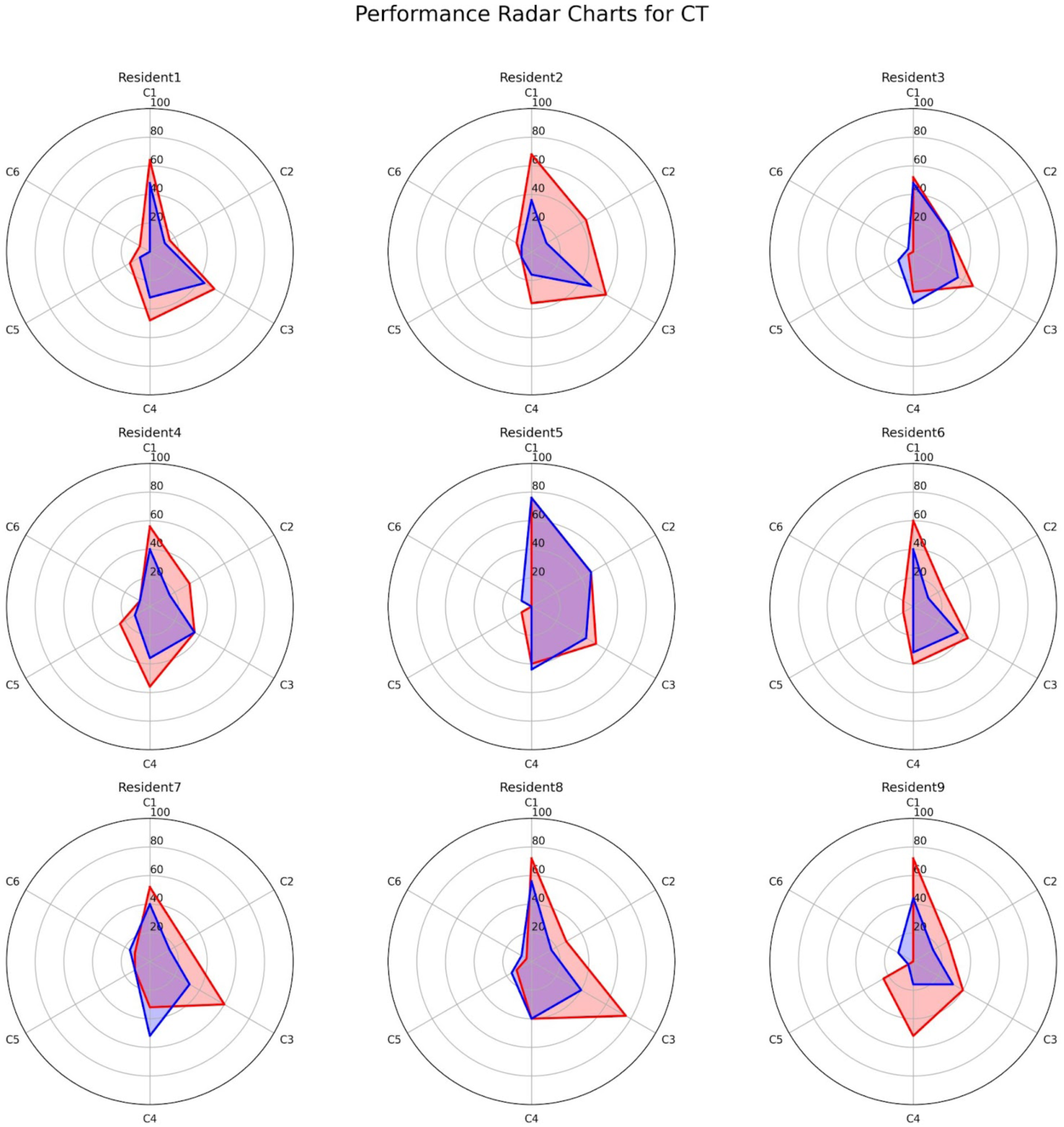
Radar chart showing the rates of CT modification for each criterion (C1–6) for nine first-year residents. Red and blue indicate the modification rates of the first and last terms, respectively. Criteria 1. Addition of missing positive findings: criterion 2. Deletion of Findings: Criterion 3. Addition of negative findings: criterion 4. Correction of the expression of findings: Criterion 5. Correction of interpretation of findings: Criterion 6. Proposals for additional tests and treatments

**Supplemental Figure 3.**
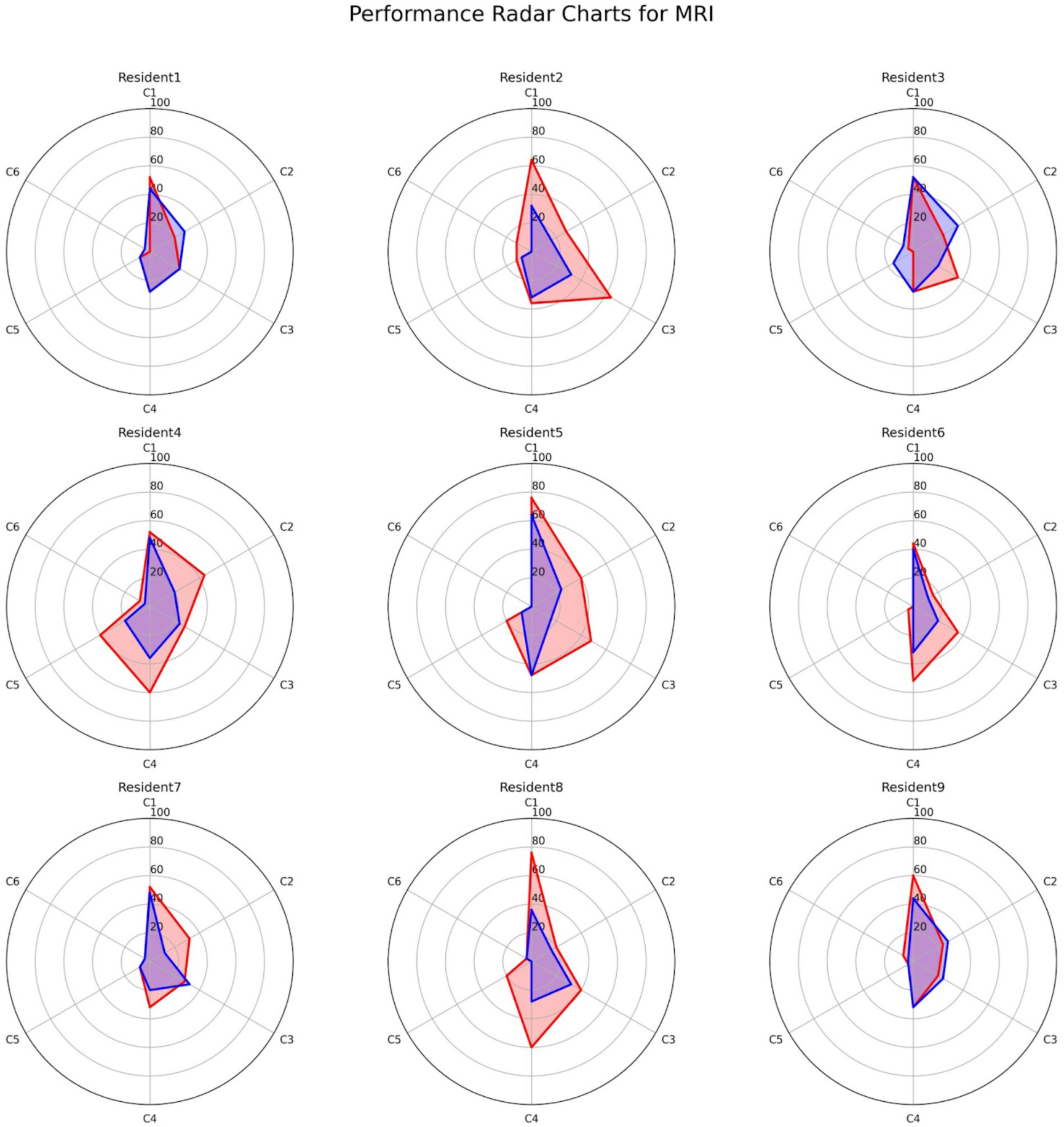
Radar chart showing the rates of MRI modification for each criterion (C1–6) for nine first-year residents. Red and blue indicate the modification rates of the first and last terms, respectively. Criteria 1. Addition of missing positive findings: criterion 2. Deletion of Findings: Criterion 3. Addition of negative findings: criterion 4. Correction of the expression of findings: Criterion 5. Correction of interpretation of findings: Criterion 6. Proposals for additional tests and treatments

